# A direct, urine-based test to diagnose acute Lyme disease using actively secreted peptidoglycan as a biomarker

**DOI:** 10.64898/2026.05.05.26352475

**Authors:** Osamudiamen Ebohon, Saadman S. Ahmad, Jules Dressler, Brenda Y. Rosario Perez, Karl L. Ocius, Marcos M. Pires, Lise E. Nigrovic, Brandon L. Jutras

## Abstract

Lyme disease is a growing and prominent human health problem caused by a group of spirochaetal bacteria that belong to the *Borrelia* genus. Persistent Lyme disease infection produces a multi-system disorder that may result in severe arthritis, carditis, neurological problems, and even death. Preventing severe disease requires immediate treatment, but current approaches to diagnose Lyme disease are indirect, serology-based assays that may fail early in infection. All Lyme disease-causing *Borrelia* species shed distinct and unique fragments of their peptidoglycan cell wall during growth. We exploited this fundamental biological process to develop an acute, urine-based diagnostic test. Using a cocktail of unique and highly specific monoclonal antibodies, our ELISA-mediated approach accurately reports on the status of an active, acute infection, in a laboratory animal model of Lyme disease, as well as humans. This rapid, simple, and innovative approach detects an active infection in as few as 3 days of transmission and in 88% of human patients yet to seroconvert—more than ∼2 weeks before serology would be positive.

## MAIN

Lyme disease (or Lyme Borreliosis) is the most common vector-borne illness in Europe and North America (1). Five genospecies of the *B. burgdorferi* sensu lato complex—*B. burgdorferi sensu stricto* (hereafter *B. burgdorferi*)*, B. afzelii, B. garinii, B. bavariensis* and *B. spielmanii*— are pathogenic to humans and are transmitted through the bite of infected *Ixodes spp*. ticks (2,3). A recognizable early symptom of Lyme disease is an erythema migrans lesion (i.e., bullseye rash), which is often accompanied by less specific ailments, such as fatigue, fever, myalgia and joint pain (4). Prompt doxycycline treatment is often sufficient to eliminate the infection, but delays can lead to disseminated and debilitating disease that impacts the neuro-musculoskeletal and cardiac systems.

The Centers for Disease Control and Prevention (CDC) recommends a two-tiered serological test for Lyme disease diagnosis but this approach has some well-recognized limitations (5,6). For instance, detectable titers require time (3 to 6 weeks) and a functional immune system (7). Even if enough time has elapsed to develop detectable titers, the results may be ambiguous and difficult to interpret, and positive results report on exposure, not the status of the infection. These issues are compounded by the overall accuracy of the method—in patients with early Lyme disease (i.e., EM lesion alone) serology fails upwards of 60 – 70% of the time (8,9). Because timely diagnosis and treatment are critical in Lyme disease to prevent potentially severe outcomes associated with missed or delayed intervention, there is an urgent need for a simple, accurate, and rapid diagnostic test capable of detecting active infection at the earliest stages.

Direct Lyme disease diagnosis using patient tissue to culture viable spirochetes is expensive, technically challenging, and unreliable (i.e., high false-negative rate). To overcome these limitations, we sought to develop a biomarker-based assay that can accurately detect an active infection across Lyme disease-causing *Borrelia* species. This approach requires abundant, unique, and bio-available molecule(s). Recent studies by our group, and others, have demonstrated that *B. burgdorferi* sheds fragments of its peptidoglycan (PG) cell wall during growth (10–12). PG is an essential mesh-like biopolymer that is thought to account for ∼10% of the total dry weight of a gram-negative cell (13). *B. burgdorferi* PG fragments (or muropeptides) are not only abundant, but exceptionally unique in that they contain L-Ornithine (L-Orn) as the diamine in the peptide chain and glycan strands that terminate with the trisaccharide Glc*N*Ac-Glc*N*Ac-1,6 anhydroMur*N*Ac (10,12,14). The latter has only ever been described in the *Borrelia* genus. Here, we exploit these atypical features to create an ELISA-based, urine biomarker assay that detects an active infection, in mice and humans, within 72 hours of transmission.

Human Lyme disease patients naturally produce specific, anti-*B. burgdorferi* PG antibodies, as do animals injected with purified PG (12). Together, we reasoned that it would be possible to capture individual B cell clones from rabbits vaccinated with purified *B. burgdorferi* PG fragments to produce monoclonal antibodies (mAbs). Individual clones were screened for cross-reactivity to abundant muropeptides containing 1,6 anhydroMur*N*Ac (anhMur*N*Ac), which we natively purified (see methods). The anhMur*N*Ac modification is important since it acts as a universal cap; the cap must be removed to accommodate growth and thus are present in released PG (15). Reactive clones were then counter-screened using muropeptides common to gram-positive and -negative bacteria (i.e., muramyl-tripeptide containing L-Lysine (L-Lys) or meso-Diaminopimelic acid (m-DAP), respectively). Four clones were recombinantly produced and used in immunoprecipitation (IP) assays with purified muropeptides from *B. burgdorferi*, that were generated by enzymatically treating intact PG sacculi. Specific, high-affinity interactions were assessed using quantitative, LCMS analysis using the input (digested PG, dPG) as a control in mock-IP reactions. Each of the four monoclonal antibodies bound to unique features present in dPG preparations, several of which contained anhMur*N*Ac (Figs. 1A, B, and S1 – S6). We then developed an ELISA-based assay to detect released *B. burgdorferi* PG using all possible combinations of capture and detection pairs. Capture antibodies were affixed to microtiter plates in each instance and were unmodified, while detection antibodies were derivatized to covalently conjugate biotin for streptavidin: HRP-mediated quantification. Each permutation yielded variable results when assessed on titrations of spent *B. burgdorferi* culture media, but a single combination (capture: 2H10/2G10, detection: 1A4/1B8) produced the most sensitive detection conditions (Figs. 1C and S7). Similar dose-response performance was attained using purified *B. burgdorferi* dPG spiked into unspent culture media or in buffered conditions (Fig. 1C and D).

**Figure 1.**
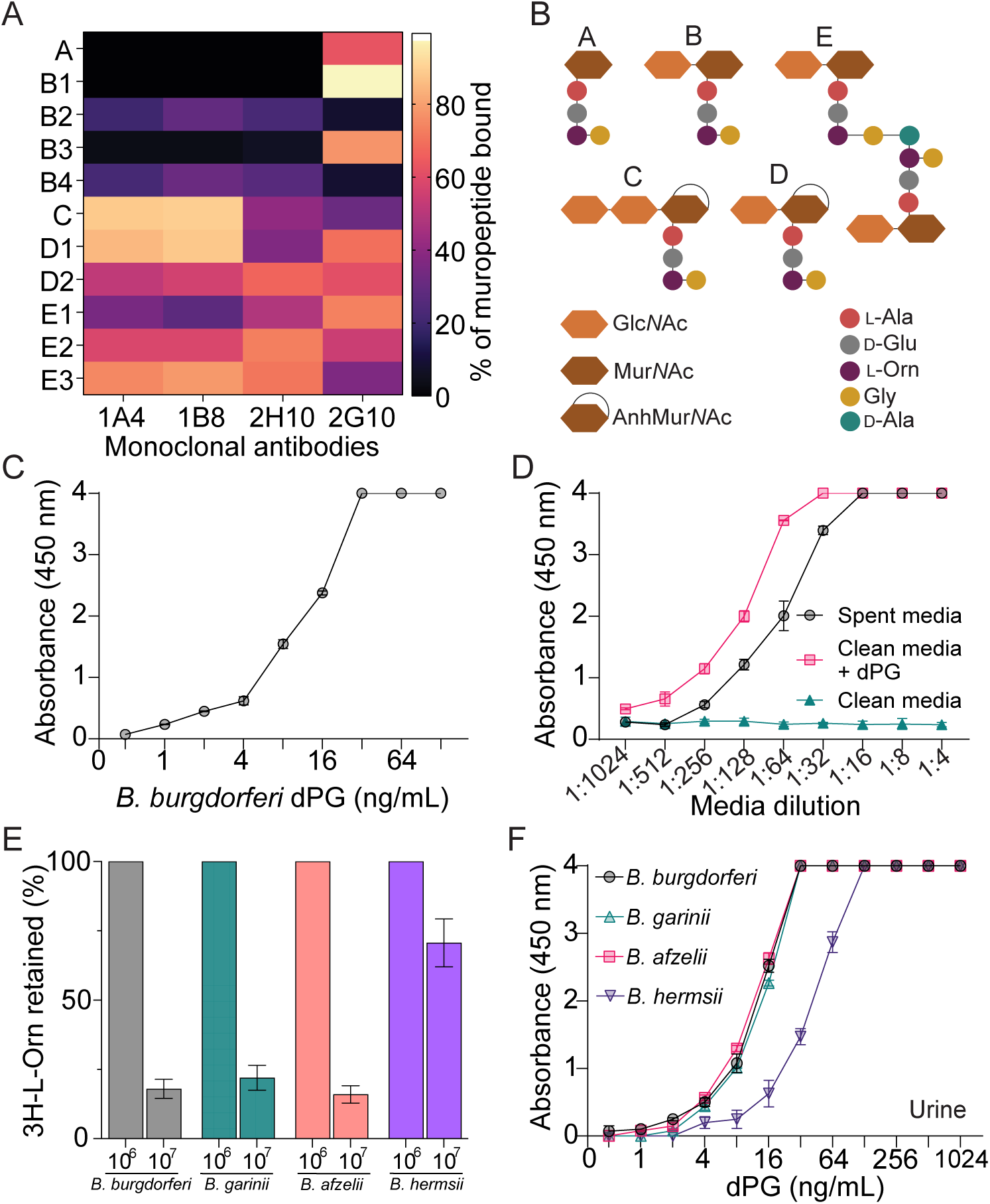
Monoclonal antibody cocktail specifically detect released *Borreliae* PG. **(A)** Each monoclonal antibody (mAb) were incubated with purified, enzyme-digested *B. burgdorferi* PG (dPG). Immunoprecipitation, coupled with quantitative LCMS, identified high-affinity interactions for four different mAb. Heat map indicates the percent of each muropeptide bound, relative to input control. **(B)** Cartoon schematic of the muropeptide structure bound by the mAbs in ‘A’. The numbers associated with each muropeptide species indicate enantomers or isobarics present in *B. burgdorferi* PG. **(C)** A cocktail of capture and detection mAb pairs is a sensitive approach to detect purified, dPG produced by *B. burgdorferi.* Values were background subtracted using the absorbance values of a PBS control. **(D)** The same cocktail of capture and detection mAb pairs provide high affinity binding to released *B. burgdorferi* PG in spent culture medium. *B. burgdorferi* were cultured to late-log, cells harvested, spent cultured media filtered (0.2 μm), and titrations were assayed by ELISA. Control reactions included sterile culture media, processed similarly, or the same sterile media spiked with dPG. **(E)** Lyme *Borrelia spp*. release PG. Each species was pulse labeled with 5uCi of 3H L-Orn for 2-3 generations, washed, and back-diluted into fresh media. Pre- and post-culture expansion, PG was purified and quantified by liquid scintillation. **(F)** The mAb cocktail cross-reacts with digested PG from different Lyme-disease causing *Borrelia* spp. Purified, dPG, attained from different *Borrelia spp.* was titrated into human urine (diluted 1:2 with PBS) and detected by ELISA. Values were background subtracted using the absorbance of a neat human urine diluted with PBS (1:2) that was not spiked with PG. Data points with absorbance of 4 were beyond the detection limit of the spectrophotometer.

The *Borrelia* genus is sub-divided in two clades that cause either Lyme disease or relapsing fever. High genotypic synteny exists throughout the group despite vast phenotypic diversity in terms of clinical presentation, geographic range of tick vector, as well as pathogen biology (16,17). With these considerations in mind, we questioned whether other *Borrelia spp.* behave like *B. burgdorferi* and release PG during growth; an essential biological feature if our biomarker assay is to perform equally across the Lyme disease group. Lyme disease-causing species *B. garinii*, *B. afzelii,* and *B. burgdorferi* (control) were compared to the relapsing fever spirochete *B. hermsii* for muropeptide release using classical pulse-chase isotope labelling studies. Each species was propagated in 3H-L-Orn containing media for 2 – 3 generations, unincorporated material was removed by washing harvested cells, and the same cells were then used to inoculate media lacking radioisotope. After culture expansion, cells were enumerated prior to extracting PG for liquid scintillation analysis. Approximately three generations of growth yielded an ∼80% reduction in 3H-L-Orn-derived signal in PG (∼40% loss per generation) in all Lyme disease-causing *Borrelia spp.*, consistent with earlier studies using *B. burgdorferi* (Fig. 1E and (12)). Curiously, *B. hermsii* retained significantly more 3H-L-Orn signal, a drastic difference from the Lyme disease clade, suggesting these bacteria may have some ability to recycle muropeptides (Fig. 1E). We also assessed the cross-reactivity of our ELISA approach using purified, dPG, isolated from each species. PG purification is a long, multi-step process that generates insoluble sacculi prior to digestion. Despite isolating PG from the same number of bacteria in each instance, the total amount of material in each preparation varied (Fig. S8A and B). We used the LCMS data collected from each to normalize preparations such that the total concentration of muropeptides fragments was equal and assayed each for cross-reactivity by ELISA (Fig. S8C and S8D). All members of the Lyme disease clade were nearly indistinguishable by ELISA using normalized material that was in buffer, or spiked into clean human urine, while relapsing fever PG preparations cross-reacted and produced signal, but to a lesser extent (Figs. 1F and S9). High-sensitivity dPG detection in urine is noteworthy, as we’ve recently discovered that polymeric PG, likely produced by dead and/or dying bacteria, persists in mice and humans, whereas fragments mimicking those released during active infection (i.e., dPG) are rapidly shed and thus could be detectable following bladder clearance (11,12).

The human body contains approximately equal numbers of bacterial and host cells (18). Bacterial replication, PG remodelling, and turnover may produce cell wall material that could interfere with a biomarker-based approach to Lyme disease diagnostics. To that end, we challenged our mAb capture-detection pairs using dPG purified from *Escherichia coli* and *Staphylococcus aureus.* Each species produces PG stem peptides with different amino acids that are representative of the dichotomy present in PG chemistry (i.e., gram-negative (m-DAP); gram-positive (L-Lys)). *B. burgdorferi* was the only Lyme disease PG sample tested in these studies since our assay performs similarly across the clade (Figs. 1F and S9). After performing the same LCMS-based concentration normalization steps described above, we found no cross-reactivity with either preparation, even when incubated at 1,000-fold higher than the detectable limits of dPG isolated from *B. burgdorferi* (Figs. 2A and S10). Human urine spiked with normalized concentrations of each preparation similarly produced no off-target binding to dPG from *E. coli* or *S. aureus* (Fig. 2B). Urinary tract infections are common and result in elevated levels of bacteria and/or bacterial products (19–21). To understand the constraints of our approach and account for other illnesses that may cause cross-reactivity, we pushed the concentration limits of spiked urine and found that our mAb cocktail has ∼7,000-15,000-fold greater affinity for *B. burgdorferi* dPG, relative to *S. aureus* or *E. coli*, further confirming the specificity of our assay (Figs. 2C and S11).

**Figure 2.**
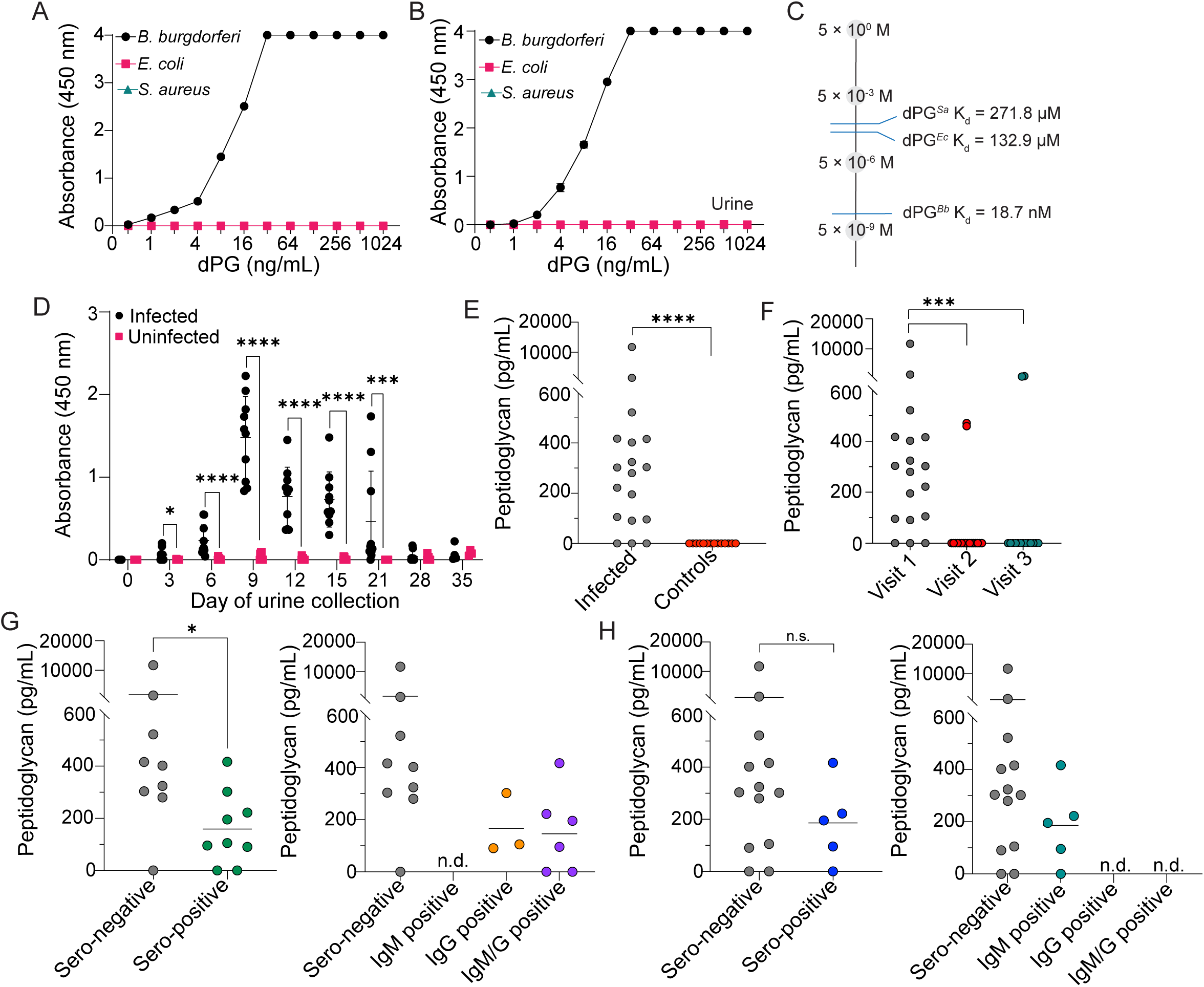
Monoclonal antibody cocktail specifically detects released PG in infected mice and humans. (**A**) ELISAs using combined capture and detection monoclonal antibody (mAb) cocktail are specific for *B. burgdorferi* PG fragments. Titrations of purified, digested PG isolated from *B. burgdorferi*, *E. coli*, and *S. aureus* were assayed by sandwich ELISA. (**B**) The same assay, described in (A), using material spiked into neat human urine. Data points with absorbance values of 4 exceeded the detection limit of the plate reader. (**C**) Apparent dissociation constants (Kd) of the mAb cocktail for dPG derived from *B. burgdorferi*, *E. coli*, and *S. aureus* spiked into human urine. Raw data are presented in Fig. S11. (**D**) Sandwich ELISA-based detection of PG in urine from *B. burgdorferi* -infected mice. Each mouse was needle inoculated subcutaneously with 100,000 *B. burgdorferi* B31-A3 cells and compared to PBS-injected animals (*n* = 10 per group). Statistical significance at each time point was determined using the Mann–Whitney U test (* *p* < 0.01; *** *p* < 0.0001). (**E**) Sandwich ELISA-based detection of PG in the urine of human Lyme disease patients confirmed to be infected by culturing skin biopsies collected at the same time as sample acquisition (visit 1). Controls were human urine samples randomly collected from endemic and non-endemic areas. *n* = 18 per group; infected: 15/18 positive (83%), controls: 0/18 (Mann–Whitney U test, **** *p* < 0.00001). Values were background subtracted using the absorbance of neat human urine diluted with PBS (1:2). (**F**) Longitudinal analysis of the same patients assayed in (E) 1 month (visit 2) or 3 months (visit 3) after doxycycline treatment. (visit 1: 15/18 PG positive, visit 2: 2/18 PG positive, visit 3: 2/15 PG positive, Mann–Whitney U test (*** *p* < 0.0001). Longitudinal data from positive patients in visits 2/3 are provided in Fig. S12. (**G**) Modified two-tier test (MTTT) results of matching serum samples for visit 1 patients (left). The same test strategy was used to determine IgM and IgG positive patients, as well as those that were both (right). Mann–Whitney U test (* *p* < 0.05), n.d., not detected. (**H**) Standard two-tier test (STTT) results for matching serum samples for visit 1 patients (left). The same test strategy was used to determine IgM and IgG positive patients, as well as those that were both (right). n.d. not detected.

Preclinical models of Lyme disease are useful since mice are a natural reservoir of *B. burgdorferi*. It is also often difficult to delineate when a human patient was bitten by an infected tick and/or the time of spirochete acquisition. We inoculated mice with the same wild-type *B. burgdorferi* B31-A3 strain used in all previous studies, collected longitudinal urine samples, and compared our results to mock-infected control animals. In as few as three days after infection, significant amounts of PG can be detected in mouse urine (Fig. 2D). PG levels peak nine days after spirochete administration, remain elevated in urine for 21 days, prior to declining to background levels (Fig. 2D). Despite the caveats of animal models and initiating infection by needle inoculation, this interval is notable since it is far too early to reliably detect seroconversion in human patients.

No FDA-approved Lyme disease diagnostic test uses urine as a matrix and early acute serum samples are often negative, which together pose a challenge for critically benchmarking our approach using validated human samples. Consequently, the only definitive way to assess the performance of our approach is to assay human urine collected from patients whose skin biopsies produced culture positive results by in vitro culture. Laboratory propagation from human skin biopsies is exceptionally challenging but it 1) confirms active infection at the time of sample collection; 2) excludes samples from patients who have received considerable therapy since antibiotic exposure frequently prevents the isolation of viable bacteria; and 3) avoids the caveats associated with patients who may have been previously exposed to Lyme disease. We were able to attain urine and serum samples from 18 different patients participating in an on-going clinical trial whose skin biopsies, acquired at the time of enrolment (visit 1), became culture positive. Longitudinal samples were collected from the same patients 4- and 12-weeks after a standard course of doxycycline therapy (visits 2 and 3, respectively). Upon enrolment (visit 1), we found that 83% of urine samples were PG positive in this cohort of properly validated Lyme disease samples (Fig. 2E). Importantly, we note that enrolment was not necessarily the patients first physician encounter; half of the patients enrolled had already received some treatment prior to visit 1, but none had finished a completed regimen (Fig. S12). When we stratified patients based on treatment, we found that PG levels were ∼4-fold higher in untreated patients, which is consistent with doxycycline mode of action as a translational inhibitor, and thus preventing the activity of lytic enzymes necessary for PG release (Fig. S12). Longitudinal analysis of urine samples after a full course of antibiotics indicated that only two patient samples remained PG positive but nearly all seroconverted (visit 2: 94%, visit 3: 93%), which strongly suggests that our approach is sensitive for patients experiencing an acute infection while being 100% specific since all negative controls yielded background signal (Figs. 2E, F, and S13). We then went back to the enrolment samples and evaluated clinical laboratory results from serum collected from the same patients during their first visit and found that 50% and 72% were negative by the modified two-tier testing (MTTT) and standard two-tier testing (STTT), respectively, thus confirming that many were at the very early stages of acute infection (Fig. 2G and H). PG levels in urine were well beyond the limit of detection in sero-negative patients (MTTT: 89%, STTT: 85%), indicating that our approach is very often positive days to weeks prior to conventional, serology-based methods (Fig. 2G and H). Collectively, our results indicate that actively released *B. burgdorferi* PG fragments represent a promising urine-based biomarker for acute Lyme disease, detectable well before seroconversion, and that our diagnostic approach rarely detects PG after treatment, in contrast to serology, which often remains positive. (Fig. 2E – H).

## DISCUSSION

Major basic and applied scientific advances have given Lyme patients hope for new preventatives and treatments, but early diagnosis has remained a challenge due to sensitivity, specificity, and temporal limitations (5,22–26). Recent serological methods, such as MTTT, have become more conclusive and capture seroconversion ∼2 – 3 weeks post-infection (25). However, they still require a competent immune system, fail at the most critical time in patient care, and will often remain positive after treatment (Fig. 2 and (8,27–30)). Direct culture and/or PCR offer alternatives that do not rely on host immune responses but are technically challenging, require 3 – 6 weeks to produce a definitive result, and/or are unreliable (27,28,31,32). Building on prior evidence that *B. burgdorferi* sheds PG fragments during growth (10,12), we demonstrate that this feature is shared by the Lyme disease-causing clade (Fig. 1) and exploit it to report the first PG-based direct diagnostic test for a bacterial infection. Our findings demonstrate that PG can be reliably detected in the urine of both infected mice and human Lyme disease patients (Fig. 2). These results provide strong conceptual and technical support for PG as a viable biomarker of an active infection that is detectable within days of spirochete transmission.

Diagnostic biomarkers play a central role in modern medicine by enabling early detection, improving clinical decision-making, and reducing reliance on invasive procedures. Ideal biomarkers are unique molecular signatures that are both abundant and readily detectable in a disease state but absent in healthy patients. Bacterial PG as a specific biomarker of acute illness is counterintuitive since it is a ubiquitous feature of virtually all bacterial cells—many of which are present on and in all humans, regardless of health status. In contrast to most gram-negative bacteria, which recycle 1,6 anhydro-containing muropeptides during growth, Lyme disease-causing spirochetes release their muropeptides during growth (Figs. 1, S7, and (10,12)). These liberated cell wall fragments are not limiting but rather constantly shed during active replication, prior to a completed course of antibiotics, and their chemical properties have unprecedented features that are unlike any other bacterium (Figs. 2E, F, S6, S7, and (10,12)). The collective consequences of each peculiarity in Lyme *Borrelia spp.* PG (e.g., L-Orn-Gly and Glc*N*Ac-Glc*N*Ac-anhMur*N*Ac muropeptides) are likely essential for monoclonal antibody specificity (Figs. 1C, 2A-B, S1 – S6 and (12,14,33)). Another important factor to the overall success of our approach is that persistent *B. burgdorferi* PG after treatment consists of polymeric PG, likely derived from dead/dying bacteria, whereas actively growing bacteria release smaller PG fragments that are cleared after treatment and/or the localized stages of disease (Fig. 2D, F – H, and (10,12)).

A limitation of our study is the relatively small number of human urine samples tested. In the absence of FDA-approved urine-based diagnostics for LD, comparative studies are not possible. We circumvented this issue by only assessing patients with culture-confirmed, acute Lyme disease, which limits our sample size. Future studies using Lyme disease biobank urine samples, collected at different stages of acute illness, will allow us to stratify different cohorts and assess both when our innovative approach succeeds and fails (34–36).

## MATERIALS AND METHODS

### Study design

This study was designed to develop a novel PG–based direct diagnostic assay for acute Lyme disease. Monoclonal antibodies (mAbs) were generated recombinantly in Chinese hamster ovary (CHO) cells and subjected to systematic screening and characterization using multiple biological and technical replicates. Peptidoglycan isolation was performed using established and previously published protocols, as detailed below. All animal experiments were conducted in C3H/HeJ mice in accordance with protocols approved by the Institutional Animal Care and Use Committee (IACUC) of the Feinberg School of Medicine, Northwestern University. Mice were purchased from The Jackson Laboratory at 4 weeks of age and acclimated for two weeks prior experiment initiation. Ethical/IRB committees of both Harvard University and Tufts University gave ethical approval for this work, as part of a NIH-funded program project grant (P01AI181934). All human patient samples were coded, and information was blinded to the scientists performing the studies.

### Bacterial strains and culture conditions

The highly infectious *Borrelia burgdorferi* B31-5A3 strain used in this study was kindly provided by Jenifer Coburn (Medical College of Wisconsin). *Borrelia garinii* (CIP 103362) and *Borrelia afzelii* (BO23) were supplied by Frank Gherardini (NIH/Rocky Mountain Labs). *Borrelia hermsii* strain HS1 was provided by Troy Bankhead (Washington State University). All *Borrelia spp.* were cultured in Barbour–Stoenner–Kelly II (BSK-II) medium (pH 7.6) supplemented with 6% heat-inactivated rabbit serum (Pel-Freez Biologicals) at 34°C without agitation until mid- to late-logarithmic growth phase (36), unless stated otherwise (see below). Prior to use, BSK-II medium was filter-sterilized using sequential 0.45 μm and 0.2 μm filters (EMD Millipore). *Escherichia coli* K-12 strain MG1655 and *Staphylococcus aureus* FDA 209 were cultured in Luria–Bertani (LB) broth at 37°C with agitation until reaching an optical density of ∼0.5, assessed using 600 nm wavelength. All bacterial cultures were harvested by centrifugation at 3,500 × g for 15 min, washed three times with phosphate-buffered saline (PBS), and stored at −20°C until further processing.

### Isotope labelling and PG release studies

Radiolabeling studies were performed essentially as previously described (12). Each *Borrelia spp.* was cultured in 40 mL of BSK-II complete media to a density of 10^7^ cells/mL. Cells were harvested by centrifugation, and gently washed with, and resuspended in minimal media (25% BSK-II, 1.2% rabbit serum, buffered with PBS) with 5 uCi/mL of 3H-L-Orn (ARC American Radiolabeled Chemicals). After ∼3 generations to pulse-label *Borrelia spp.* PG, cells were harvested, and washed three times with PBS, and once with complete BSK-II equilibrated to 34C, prior to be suspended. Each culture was enumerated and further diluted to a final concentration of 10^6^ cells/mL. One half of the culture was then removed, this fraction was split in half, and PG was purified by boiling pelleted and washed cells by boiling each in 5% SDS (final, vol/vol). Insoluble material was collected by centrifugation and starting radioactivity was determined by liquid scintillation by calculating the average (+/- SD) of both preparations. The other half of the culture was incubated at 34°C until it reached a final density of 10^7^cells/mL, prior to once again splitting in half, and repeating the procedure described above. Any differences in final cell density between species were used to normalize the liquid scintillation values attained.

### Peptidoglycan (PG) isolation and processing

PG was isolated using a modified version of previously described protocols (14,38), adapted from the classic method of Glauner (39). Briefly, *Borrelia* and *E. coli* cells were resuspended in cold PBS, whereas *S. aureus* cells were resuspended and subjected to repeated sonication cycles (60 s sonication followed by 60 s on ice). Cell suspensions were added dropwise to boiling 10% (w/v) SDS to achieve a final SDS concentration of 5%. Samples were boiled for 1 h and subsequently washed five times with warm (∼40°C) nanopure water. PG sacculi were pelleted by ultracentrifugation at 218,300 × *g* for 60 min at 30°C. For *Borrelia* PG sacculi, samples were treated with chymotrypsin (0.3 mg/mL) overnight at 37°C with shaking at 540 rpm. In contrast, *E. coli* and *S. aureus* PG preparations were sequentially treated with lipase (1 mg/mL, 3 h), benzonase nuclease (4 μL/mL, 2 h), and chymotrypsin (0.3 mg/mL, overnight). Following enzymatic digestion, SDS was added to a final concentration of 0.5% (v/v), and samples were heat-inactivated at 80°C for 30 min, washed three additional times, and pelleted as described above. For *S. aureus*, an additional acid hydrolysis step was performed by incubating PG sacculi in 1 M HCl for 48 h at 4°C with rotation, followed by two washes and ultracentrifugation.

Muropeptides were generated by digesting PG sacculi with mutanolysin (400 U/mL final concentration) in 5 mM NaHPO_4_/NaH_2_PO_4_ buffer (pH 5.5) overnight at 37°C with shaking at 540 rpm. An additional 400 U of mutanoylsin was added the following day and digested for another 24 h. The enzyme was then inactivated (100°C for 10 min), insoluble material was removed by centrifugation at 22,000 × *g* for 30 min, and soluble muropeptides were transferred to pre-weighed tubes. Soluble muropeptides were then lyophilized and quantified using an analytical balance prior to secondary methods (see text and below).

### Muropeptide processing for LCMS analysis and normalization

Lyophilized muropeptides were dissolved in 150 μL of saturated sodium borate buffer adjusted to pH 9.25. Reduction was initiated by the gradual addition of 50 μL sodium borohydride (100 mg/mL) and allowed to proceed for 1 h. The reaction was quenched by acidification with approximately 10 μL formic acid to pH 3. Samples were immediately frozen and dried under high vacuum. The resulting residue was reconstituted in 200 μL of a 9:1 (v/v) mixture of water and acetonitrile containing 0.1% formic acid, followed by 10 min of bath sonication. Insoluble debris was removed by centrifugation at 13,000 × *g* for 10 min at 4°C, and 180 μL of the clarified extract was transferred to LCMS vials for subsequent analysis. LCMS analyses were performed as previously described (12,38) on a Shimadzu LCMS9030 QToF system coupled to a LC-40B X3 UPLC, a SIL-40C X3 autosampler (10°C), and a CTO-40C column oven (40°C). Muropeptides from *Borrelia* genospecies and other bacterial clades used for downstream analysis were normalized based on total chromatographic peak area. Individual muropeptide species were identified based on accurate mass and retention time, and peak areas were integrated using the same processing parameters for all samples. The total muropeptide peak area for each sample was calculated by summing the areas of all detected muropeptide peaks and was used to normalize peptidoglycan abundance across samples prior to downstream analysis.

### Monoclonal antibodies production

Monoclonal antibodies (mAbs) were generated through Abclonal using purified mutanolysin-digested muropeptides and intact PG sacculi as immunogens. Abclonal was responsible for immunizations and initial B cell screening. Additional screening, as well as counter-selection were performed in house (see below). To begin, both forms of PG (digested and intact, 200 μg each) were used to immunize two New Zealand White rabbits, followed by monthly booster injections (200 μg) over a period of seven months. After confirmation of robust antibody responses, animals were euthanized, splenocytes were harvested, and B cell clones were generated and screened using Abclonal’s proprietary methods. Approximately 500 B cell clones were screened for binding to distinct biotinylated *B. burgdorferi* muropeptides (see below), resulting in the identification of 93 clones producing mAbs with high affinity for *B. burgdorferi* PG. These clones were then counter-screened using commercially available muramyl-dipeptide (MDP), muramyl-tripeptide (MTP) containing either m-DAP, or L-Lys, which are all available through Invivogen. All three muropeptides were biotinylated using methods described below. The counter screen yielded 17 specific antibodies, which were further evaluated and prioritized based on their affinities. Four mAbs exhibiting high specificity and affinity for *B. burgdorferi* PG were selected. The heavy and light chains were cloned into a proprietary TurboCHO viral expression vector and recombinantly expressed in CHO cells by GenScript. Recombinant mAbs were purified and used for all subsequent experiments in this study.

### Native *B. burgdorferi* muropeptide purification

PG isolated from 10 L cultures of *B. burgdorferi* in complete BSK-II medium was isolated and purified as described above. To isolate specific muropeptides for monoclonal antibody screens, we coupled size exclusion chromatography (SEC) with preparative liquid chromatography (prepLC). Bio-Gel P2 beads (Bio-Rad) were rehydrated overnight with LCMS grade H_2_O: 95% 200 proof EtOH (9:1 v:v) and packed in a 1 x 20 cm column filled with SEC solver. Fractions (0.5 – 1.0 mL) were collected and assessed for the presence of muropeptides using a DeNovix spectrophotometer (at 205 and 215 nm wavelengths).

Fractions containing muropeptides were then fractionated using a Shimadzu LC-40D HPLC, a SIL-40C autosampler (10°C) and a CTO-40C column oven (40°C) coupled to an FRC-40 fraction collector, alternating between using a Hypercarb column (4.6 mm x 100 mm, 5 μm particle size; Thermo Scientific) and a C18 column (4.6 mm x 250 mm, 5 μm particle size; Restek) to optimize resolution. Solvent A (0.1% formic acid in water) and solvent B (0.1% formic acid in MeCN) at a constant flow rate of 0.5 mL/min. Initial conditions were 5% B for 3 min before increasing the gradient to 20% B at 10 min. The gradient was ramped up to 95% B at 12 min and held for 2 min. This was decreased back down to 5% at 15 min and held till the end of the run at 20 min. Flow injection analysis (FIA) MS, bypassing the use of a separation column allowed for high throughput analysis of each fraction. An isocratic gradient of 50% A (0.1% formic acid in water) and 50% B (0.1% formic acid in MeCN) at a flow rate of 0.2 mL/min for 2 min was utilized. Fractions were analyzed on a Shimadzu LCMS9030 QToF system coupled to a LC-40B X3 UPLC, and a SIL-40C X3 autosampler (10°C). Data was collected data in MS mode only, from 400 m/z to 2,000 m/z. In case of contaminating muropeptides, further separation was achieved with SEC.

### Biotinylation of muropeptides for antibody screen and counter-screen

Isolated and purified Glc*N*Ac-AnhMur*N*Ac-L-Ala-D-Glu-L-Orn-Gly (both isoforms), Glc*N*Ac-Glc*N*Ac-Mur*N*AcAnh-L-Ala-D-Glu-L-Orn-Gly, and commercial muropeptides described above were biotinylated using EZ-Link PFP-Biotin (Thermo Scientific) as per the manufacturer’s recommended conditions. Unreacted biotin was removed using SEC as outlined above.

### Epitope characterization of monoclonal antibodies

Each mAb (200 µg/mL) was incubated with mutanolysin-digested *B. burgdorferi* PG (20 µg/mL) overnight at 4°C. Unbound PG was separated from mAb–PG complexes using centrifugal filter units with a 10-kDa molecular weight cutoff. The retained mAb–PG complexes were washed five times with 200 µL PBS, and the combined flowthrough fractions were pooled and analyzed as the unbound PG fraction. PG–mAb complexes were disassociated by boiling and mixture was re-applied to fresh centrifugation columns to separate liberated muropeptides from each mAb. Input PG, unbound flowthrough, and bound (retentate) fractions were analyzed by LCMS as described above. Muropeptide species were identified based on accurate mass and retention time, and retention efficiency was quantified as the percentage of each muropeptide peak area recovered in the retentate relative to the corresponding input All data from these assays are available in supplementary information.

### Monoclonal antibody sensitivity and specificity

Sandwich ELISAs were used to determine the specificity and sensitivity of the mAbs for *B. burgdorferi* PG as described previously (11). Briefly, ELISA plates were coated overnight at 4 °C with mAbs (75 µL/well; 10 µg/mL in 0.05 M carbonate–bicarbonate buffer, pH 9.6), washed with PBS–T (0.05% Tween-20), and blocked for 1 h at 37°C with fish serum blocking buffer. Plates were incubated for 1 h at 37°C with serial dilutions of digested PGs from *Borrelia* genospecies, *E. coli*, *S. aureus* or spent culture media from *B. burgdorferi,* washed and incubated with biotinylated mAb (2 µg/mL) for 1 h at 37°C. Following incubation with HRP-conjugated streptavidin (1:10,000), the signal was developed using TMB substrate, stopped with 1.5 N H₂SO₄, and absorbance measured at 450 nm.

Assay sensitivity was enhanced by screening mAb cocktails using pairwise combinations of two capture mAbs (mixed 1:1 at 10 µg/mL) and two biotinylated detection mAbs (mixed 1:1 at 2 µg/mL). The optimal mAb cocktail was selected and used for all subsequent experiments. To account for matrix effects, PG was spiked into pooled human urine (Innovative Research, IRHUURE50ML–53380) diluted 1:2 and analyzed using the sandwich ELISA described above.

To determine the dissociation constant (K_d_) of the mAb cocktail for each type of PG by ELISA, total muropeptide profile concentrations were first converted to molar units, followed by preparation of serial dilutions from a saturating concentration. Digested PG (2 µL; 400 µg/mL, 0.8 µg total) was analyzed by LCMS. Muropeptides were identified by accurate mass, and theoretical molecular weights were assigned based on structure. Relative abundances were determined from integrated peak areas, and the average PG molecular weight was calculated as a peak-area–weighted mean. For each identified muropeptide, the theoretical molecular weight was multiplied by its corresponding LCMS peak area, and the sum of these products was divided by the total peak area of all detected muropeptides. PG concentrations were then converted to molar units based on the calculated average molecular weight. Serial dilutions of PG from *B. burgdorferi*, *E. coli*, and *S. aureus* were prepared, and 75 µl of each dilution was added per well. *B. burgdorferi* PG was diluted starting from 0.1 µM, whereas *E. coli* and *S. aureus* PG were diluted starting from 2 mM. All subsequent steps following PG addition were performed as described above.

### Assessing spent culture media for the presence of released muropeptides

A sandwich ELISA was used to detect released peptidoglycan (PG) fragments (muropeptides) in spent culture media. Plates were coated with a monoclonal antibody (mAb) cocktail and blocked as described above. *B. burgdorferi* strain B31-5A3 was grown in BSK-II medium to late mid-log phase, and cultures were centrifuged at 3,500 × g for 15 min to collect the spent culture supernatant. Neat BSK-II medium served as a negative control, while BSK-II medium spiked with digested *B. burgdorferi* PG (1 µg/mL) served as a positive control. Plates were incubated with serial dilutions of spent culture supernatant and control samples for 1 h at 37°C. Wells were then incubated sequentially with a biotinylated detection mAb cocktail, HRP-conjugated streptavidin, and TMB substrate. The reaction was stopped with 1.5 N H₂SO₄, and absorbance was measured at 450 nm.

### Peptidoglycan detection in *B. burgdorferi-*infected mice

Four-week-old female mice were obtained from The Jackson Laboratory and housed five per cage under a 12-h light/dark cycle with ad libitum access to standard chow and water. *B. burgdorferi* cultures were grown to mid-log phase (5 ×10^7^ cells/mL), washed three times with cold sterile PBS, and counted manually in triplicate using a hemocytometer. Cells were resuspended to 10^6^ cells/mL in PBS, and mice were inoculated subcutaneously with 100 µL (10^5^ cells) using a 27-gauge, 0.5-inch needle. Urine was collected every 3 days for the first 14 days postinfection and weekly thereafter up to 35 days by manual compression of the abdominal wall (40). Urine samples were diluted 1:2 and analyzed for PG using the sandwich ELISA described above with biotinylated mAb at 3 µg/mL (equimolar amount of two biotinylated detection mAbs at 3 µg/mL was used).

### Peptidoglycan detection in Lyme disease patients

Urine samples were collected from patients with acute Lyme disease at the initial clinic visit and at follow-up visits at 4 and 12 weeks after a standard course of doxycycline treatment. During the first visit, a skin biopsy at was obtained adjacent to the leading edge of a suspected bullseye rash and used to inoculate BSK-II complete media. Coded, experimental samples (i.e., urine and serum) analyzed in this study were only included if skin biopsies were culture positive. Urine from individuals confirmed to be Lyme disease–negative served as controls. Negative control samples were attained from both endemic and non-endemic regions and included an approximately equal number of male and females. PG in urine was quantified using a modified sandwich ELISA as described above. Briefly, plates were coated overnight at 4°C with two capture monoclonal antibodies (mAbs) mixed at equimolar concentrations (total 10 µg/mL; 100 µL/well), followed by washing with PBS–T (0.05% Tween-20), and blocked for 90 min at room temperature (RT) with 1% BSA. Urine samples, diluted 1:2 in PBS, were added and incubated for 3 h at RT. A standard curve was generated by spiking digested *B. burgdorferi* PG into neat human urine diluted 1:2 in PBS, while neat human urine diluted 1:2 in PBS served as the blank for background subtraction; both were incubated under identical conditions. After washing, plates were incubated with biotinylated detection mAbs (two mAbs at equimolar amounts, 5 µg/mL) for 90 min at RT, followed by HRP-conjugated streptavidin (1:10,000) for 45 mins at RT. Signal was developed using Abcam high-sensitivity TMB substrate, stopped with 1.5 N H₂SO₄, and absorbance was measured at 450 nm. Blank absorbance values were subtracted from all readings, and the limit of detection (LOD) was defined as 3X the standard deviation of the blank. Each sample was analyzed in technical triplicates per assay. The experiment was independently repeated once under the same conditions. For each sample, the mean value from each run was calculated, and the final reported value represents the average of the two independent experiments.

### Clinical laboratory serology testing of acute Lyme disease patient samples

All available serum samples were tested using STTT and MTTT for IgM, IgG, IgG/IgM using the DiaSorin LIAISON Lyme IgM, LIAISON Lyme IgG, and LIAISON Lyme Total Antibody Plus kits, respectively. Tests were performed blindly at the Clinical Microbiology Laboratory at Massachusetts General Hospital.

### Statistical analysis

All statistical analyses are described in the text and figure legends and were performed using GraphPad Prism Version 10.5.0 (774).

## Data Availability

All data produced in the present work are contained in the manuscript

## Acknowledgements

This work was supported by The Assistant Secretary of Defense for Health Affairs through the Tick-Borne Disease Research Program, endorsed by the Department of Defense under Award No. HT9425-23-1-1042, which was granted to BLJ and MMP. Opinions, interpretations, conclusions and recommendations are those of the author and are not necessarily endorsed by the Department of Defense. Antibody creation and preclinical studies were supported by the Bay Area Lyme Foundation and the LymeX accelerator prize (HHS), respectively, both awarded to BLJ. Acute Lyme disease patient samples were collected through studies supported by National Institutes of Allergy and Infectious Disease (P01AI181934); granted to BLJ and other members of the P01 research group. This mechanism also supports BRP. Conceptual and technical lessons learned as a result of other sponsored research (R01AI178711, R01AI173256, R21AI159800) to BLJ, helped shape the final design of our assay. They also partially supported JD. SSA was supported by the Global Lyme Alliance and the Ryan Family Accelerator Research Fund (through the Feinberg School of Medicine). We would like to thank P01AI181934 team members John Branda and Karina Yee (Mass General Hospital/Harvard University) and Linden Hu (Tufts University) for curated patient sample data, as well as thoughtful discussion during manuscript preparation. We would also like to thank Liz Horn (Bay Area Lyme BioBank) for providing samples for method optimization, and members of the Jutras lab; Kaaren Jensen; and Kristin Rose Jutras for their thoughtful review of this manuscript.

## References

1. Mead PS. Epidemiology of Lyme disease. Infect Dis Clin North Am [Internet]. 2015;29(2):187–210. Available from: 10.1016/j.idc.2015.02.010

2. Stanek G, Reiter M. The expanding Lyme Borrelia complex — clinical significance of genomic species? Eur Soc Clin Infect Dis [Internet]. 2011;17(4):487–93. Available from: 10.1111/j.1469-0691.2011.03492.x

3. O’Connell S, Wolfs T. Lyme Borreliosis. Pediatr Infect Dis J. 2014;33(4):407–9.

4. Wormser GP. Early Lyme disease. N Engl J Med. 2006;354(26):2794–801.

5. Branda JA, Steere AC. Laboratory diagnosis of Lyme Borreliosis. Clin Microbiol Rev. 2021;34(2):1–45.

6. Centers for Disease Control and Prevention (CDC). MMWR Morb Mortal Wkly Rep. 1995 [cited 2023 Mar 23]. p. 11;44(31):590-1 Recommendations for test performance and interpretation from the Second National Conference on Serologic Diagnosis of Lyme Disease. Available from: https://www.cdc.gov/mmwr/preview/mmwrhtml/00038469.htm

7. Aguero-Rosenfeld ME, Nowakowski J, Bittker S, Cooper D, Nadelman RB, Wormser GP. Evolution of the serologic response to Borrelia burgdorferi in treated patients with culture-confirmed erythema migrans. J Clin Microbiol. 1996;34(1):1–9.

8. Horn EJ, Menefee B, Schotthoefer AM, Dempsey G, Mcardle M, Weber AF, et al. Evaluation of standard and modified two-tiered testing algorithms using well-characterized early Lyme disease samples. J Clin Microbiol. 2026;1–13.

9. Waddell LA, Greig J, Mascarenhas M, Harding S, Lindsay R, Ogden N. The accuracy of diagnostic tests for Lyme disease in humans, a systematic review and meta-analysis of North American research. PLoS One. 2016;11(12):1–23.

10. Mccausland JW, Kloos ZA, Irnov I, Sonnert ND, Grimes CL, Jacobs-wagner C. Bacterial and host enzymes modulate the pro-inflammatory response elicited by the peptidoglycan of Lyme disease agent Borrelia burgdorferi. PLoS Pathog [Internet]. 2025;21(7):1–31. Available from: 10.1371/journal.ppat.1013324

11. Mcclune ME, Ebohon O, Dressler JM, Davis MM, Tupik JD, Lochhead RB, et al. The peptidoglycan of Borrelia burgdorferi can persist in discrete tissues and cause systemic responses consistent with chronic illness. Sci Transl Med. 2025;2955(April):1–16.

12. Jutras BL, Lochhead RB, Kloos ZA, Biboy J, Strle K, Booth CJ, et al. Borrelia burgdorferi peptidoglycan is a persistent antigen in patients with Lyme arthritis. Proc Natl Acad Sci U S A. 2019;116(27):13498–507.

13. de Pedro MA, Cava F. Structural constraints and dynamics of bacterial cell wall architecture. Front Microbiol. 2015;6(May):1–10.

14. DeHart TG, Kushelman MR, Hildreth SB, Helm RF, Jutras BL. The unusual cell wall of the Lyme disease spirochaete Borrelia burgdorferi is shaped by a tick sugar. Nat Microbiol. 2021;6(12):1583–92.

15. Gilmore MC, Yadav AK, Espaillat A, Gust AA, Williams MA, Brown PJB, et al. A peptidoglycan N-deacetylase specific for anhydroMurNAc chain termini in Agrobacterium tumefaciens. J Biol Chem [Internet]. 2024;300(2):105611. Available from: 10.1016/j.jbc.2023.105611

16. Steere AC, Strle F, Wormser GP, Hu LT, Branda JA, Li X, et al. Lyme borreliosis. Nat Rev Dis Prim. 2016;2(1):1–19.

17. Koutantou M, Drancourt M, Angelakis E. Prevalence of Lyme disease and relapsing fever Borrelia spp. in vectors, animals, and humans within a one health approach in mediterranean countries. Pathogens. 2024;13(6):1–53.

18. Sender R, Fuchs S, Milo R. Revised estimates for the number of human and bacteria cells in the body. PLoS Biol. 2016;14(8):1–14.

19. Foxman B, Brown P. Epidemiology of urinary tract infections transmission and risk factors, incidence, and costs. Infect Dis Clin North Am. 2003;17:227–41.

20. He Y, Zhao J, Wang L, Han C, Yan R, Zhu P, et al. Epidemiological trends and predictions of urinary tract infections in the global burden of disease study 2021. Sci Rep. 2025;15(1):1–12.

21. Olin SJ, Bartges JW. Urinary tract infections: treatment/comparative therapeutics. Vet Clin Small Anim. 2015;45:721–746.

22. Branda JA, Strle K, Nigrovic LE, Lantos PM, Lepore TJ, Damle NS, et al. Evaluation of Modified 2-Tiered Serodiagnostic Testing algorithms for early Lyme disease. Clin Infect Dis. 2017;64(8):1074–80.

23. Gabby ME, Bandara A, Outrata LM, Ebohon O, Ahmad SS, Dressler JM, et al. A high-resolution screen identifies a preexisting lactam that specifically treats Lyme disease in mice. Sci Transl Med. 2025;795(17):1–14.

24. Simon R, Lamberth E, Stark JH, Skinner JM, Simon R, Lamberth E, et al. Expert Review of Vaccines A human Lyme disease vaccine : two steps forward on the path to prevention. Expert Rev Vaccines [Internet]. 2026;25(1). Available from: 10.1080/14760584.2025.2607482

25. Wagner L, Obersriebnig M, Kadlecek V, Hochreiter R, Ghadge SK, Larcher-senn J, et al. Immunogenicity and safety of different immunisation schedules of the VLA15 Lyme borreliosis vaccine candidate in adults, adolescents, and children: a randomised, observer-blind, placebo-controlled, phase 2 trial. Lancet Infect Dis. 2025;25:986–99.

26. You S, Shrestha M, Bourgeois J, Clendenen L, Leimer N, Lewis K, et al. Hygromycin A treatment of Borrelia burgdorferi-infected Peromyscus leucopus suggests potential as a reservoir-targeted antibiotic. J Infect Dis. 2026;233(1):96–100.

27. Feder HM, Gerber MA, Luger SW, Ryan RW. Persistence of serum antibodies to Borrelia burgdorferi in patients treated for Lyme disease. Clin Infect Dis. 1992;15(5):788–93.

28. Bil-Lula I, Matuszek P, Pfeiffer T, Woźniak M. Lyme borreliosis - The utility of improved real-time PCR assay in the detection of Borrelia burgdorferi infections. Adv Clin Exp Med. 2015;24(4):663–70.

29. Kannian P, Mchugh G, Johnson BJB, Bacon RM, Glickstein LJ, Steere AC. Antibody Responses to Borrelia burgdorferi in patients with non–antibiotic-treated Lyme arthritis. Arthritis Rheum. 2007;56(12):4216–25.

30. Peltomaa M, Mchugh G, Steere AC. Persistence of the antibody response to the VlsE sixth invariant region (IR 6) peptide of Borrelia burgdorferi after successful antibiotic treatment of Lyme disease. J Infect Dis. 2003;187(8):1178–86.

31. Niemz A, Ferguson TM, Boyle DS. Point-of-care nucleic acid testing for infectious diseases. Trends Biotechnol [Internet]. 2011;29(5):240–50. Available from: 10.1016/j.tibtech.2011.01.007

32. Tenover FC. The role for rapid molecular diagnostic tests for infectious diseases in precision medicine. Expert Rev Precis Med Drug Dev [Internet]. 2018;3(1):69–77. Available from: 10.1080/23808993.2018.1425611

33. Ahmad SS, Ebohon O, Mcclune ME, Trimble RN, Kellogg CN, Booth CJ, et al. Peptidoglycan architecture dictates protein interactions, tissue tropism, and arthritis in the Lyme disease spirochete Borrelia burgdorferi. PLoS Pathog [Internet]. 2026;22(1):1–23. Available from: 10.1371/journal.ppat.1013849

34. Horn EJ, Dempsey G, Schotthoefer AM, Prisco UL, Mcardle M, Gervasi SS, et al. The Lyme Disease Biobank: Characterization of 550 patient and control samples from the East Coast and Upper Midwest of the United States. J Clin Microbiol. 2020;58(6):1–12.

35. Levin AE, Wormser GP, Horn EJ, Karaseva N, Miller D, Kellogg H. A novel single-tier serologic test to diagnose all stages of Lyme disease. J Clin Microbiol. 2025;63(9):1–13.

36. Nigrovic LE, Neville DN, Balamuth F, Levas MN, Bennett JE, Kharbanda AB, et al. Pediatric Lyme Disease Biobank, United States, 2015–2020. Emerg Infect Dis. 2020;26(12):3099–101.

37. Barbour AG. Isolation and cultivation of Lyme disease spirochetes. Yale J Biol Med. 1984;57(4):521–5.

38. Jutras BL, Scott M, Parry B, Biboy J, Gray J, Vollmer W, et al. Lyme disease and relapsing fever Borrelia elongate through zones of peptidoglycan synthesis that mark division sites of daughter cells. Proc Natl Acad Sci U S A. 2016;113(33):9162–70.

39. Glauner J. Separation and quantification of muropeptides with High-Performance Liquid Chromatography. Anal Biochem. 1988;172:451–64.

40. Kadekawa K, Yoshimura N, Majima T, Wada N, Shimizu T, Birder LA, et al. Characterization of bladder and external urethral activity in mice with or without spinal cord injury—a comparison study with rats. Am J Physiol Regul Integr Comp Physiol. 2016;310(8):752–8.

