## Supplemental Material for "A direct, urine-based test to diagnose acute Lyme disease using actively secreted peptidoglycan as a biomarker"

Ebohon et al.

**Table of contents** **Page**

Figure S1: Structures of muropeptides enriched by monoclonal antibody affinity capture……………………………………………………………………………… 1

Figure S2: LCMS analysis of muropeptides bound to mAb 2G10…………………… 2

Figure S3: LCMS analysis of muropeptides bound to mAb 1A4……………………… 3

Figure S4: LCMS analysis of muropeptides bound to mAb 2H10……………………. 4

Figure S5: LCMS analysis of muropeptides bound to mAb 1B8……………………… 5

Figure S6: Muropeptides with a 1,6 anhydromuramic acid group that were identified by LCMS after antibody pull-down………………………………………………………… 6

Figure S7: Optimization of mAb cocktail to detect released peptidoglycan in *B. burgdorferi* spent culture medium……………………………………………………. 7

Figure S8: LCMS-based normalization of *Borrelia spp*. muropeptides…………………. 8

Figure S9: Optimized mAb cocktail cross-reacts with digested PG from different Lyme-disease causing *Borrelia spp*………………………………………………………. 9

Figure S10: LCMS-based normalization of muropeptides across different bacterial *spp.* 10

Figure S11: Disassociation constant calculations for mAb capture/detection pair………. 11

Figure S12: Antibiotic treatment status of patients relative to PG levels in urine ……….. 12

Figure S13: Quantifying *B. burgdorferi* peptidoglycan concentration in the urine of human Lyme disease patients………………………………………………………………. 13

Structure A

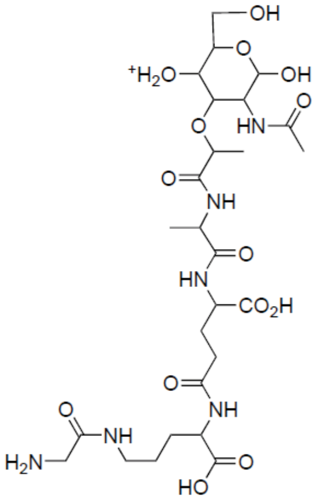

Chemical Formula: $C_{26}H_{45}N_6O_{14}^+$
Exact Mass: 665.2988

Structure B

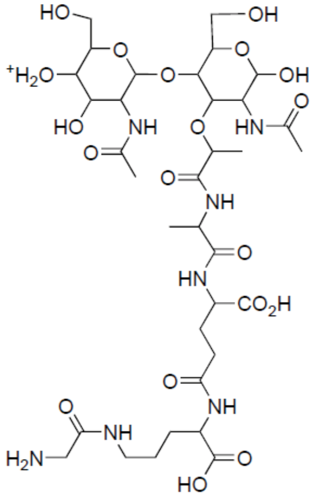

Chemical Formula: $C_{34}H_{58}N_7O_{19}^+$
Exact Mass: 868.3782

Structure C

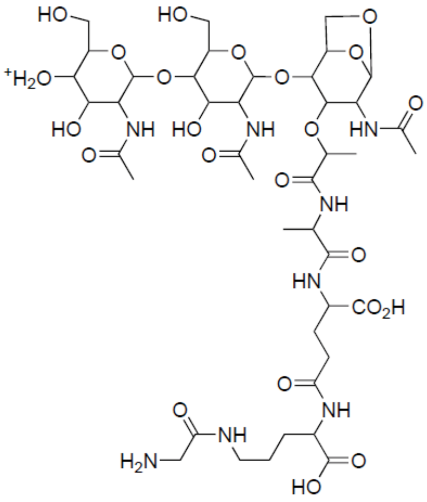

Chemical Formula: $C_{42}H_{69}N_8O_{23}^+$
Exact Mass: 1053.4470

Structure D

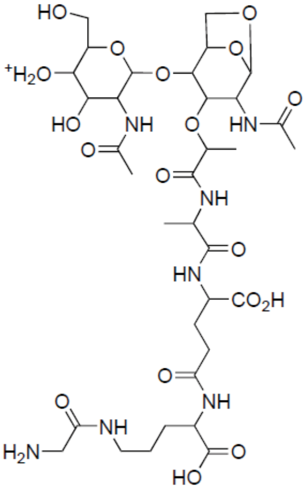

Chemical Formula: $C_{34}H_{56}N_7O_{18}^+$
Exact Mass: 850.3676

Structure E

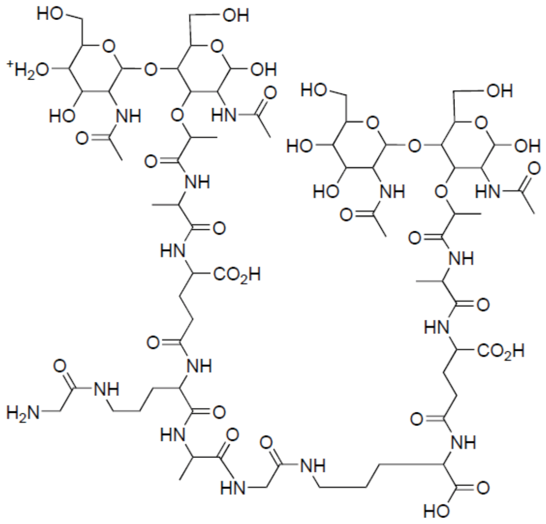

Chemical Formula: $C_{71}H_{118}N_{15}O_{38}^+$
Exact Mass: 1788.7757

**Fig. S1: Structures of muropeptides enriched by monoclonal antibody affinity capture.** Muropeptide structures were elucidated using MS-derived mass/charge data. Note that structures are predicted, based on MS data (see Figs. S2-S5), and retention time differences are due to sugar enatomers (i.e., not reduced) and the presence of isobaric structures in *B. burgdorferi* PG.

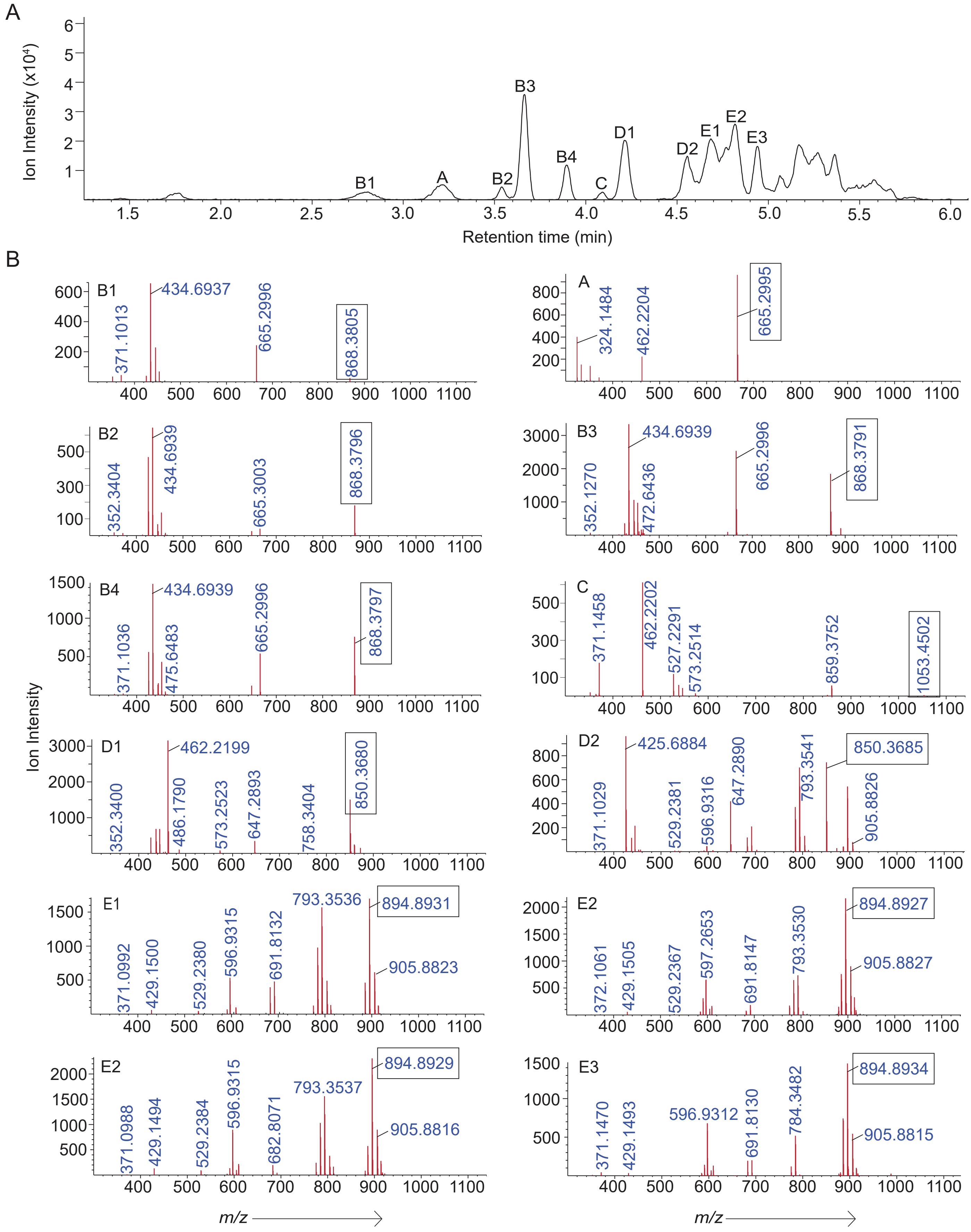


**Fig. S2: LCMS analysis of muropeptides bound to mAb 2G10.** (**A**) LCMS chromatogram showing distinct peaks corresponding to muropeptides captured by 2G10 following incubation with dPG. (**B**) Mass spectra corresponding to each chromatographic peak, confirming the identity of the bound muropeptides. The boxed region highlights the mass spectra peak at the indicated m/z corresponding to the assigned muropeptide. Crosslinked species (structure E), are detected as doubly charged ions (2+), with molecular mass approximated as 2 × m/z.

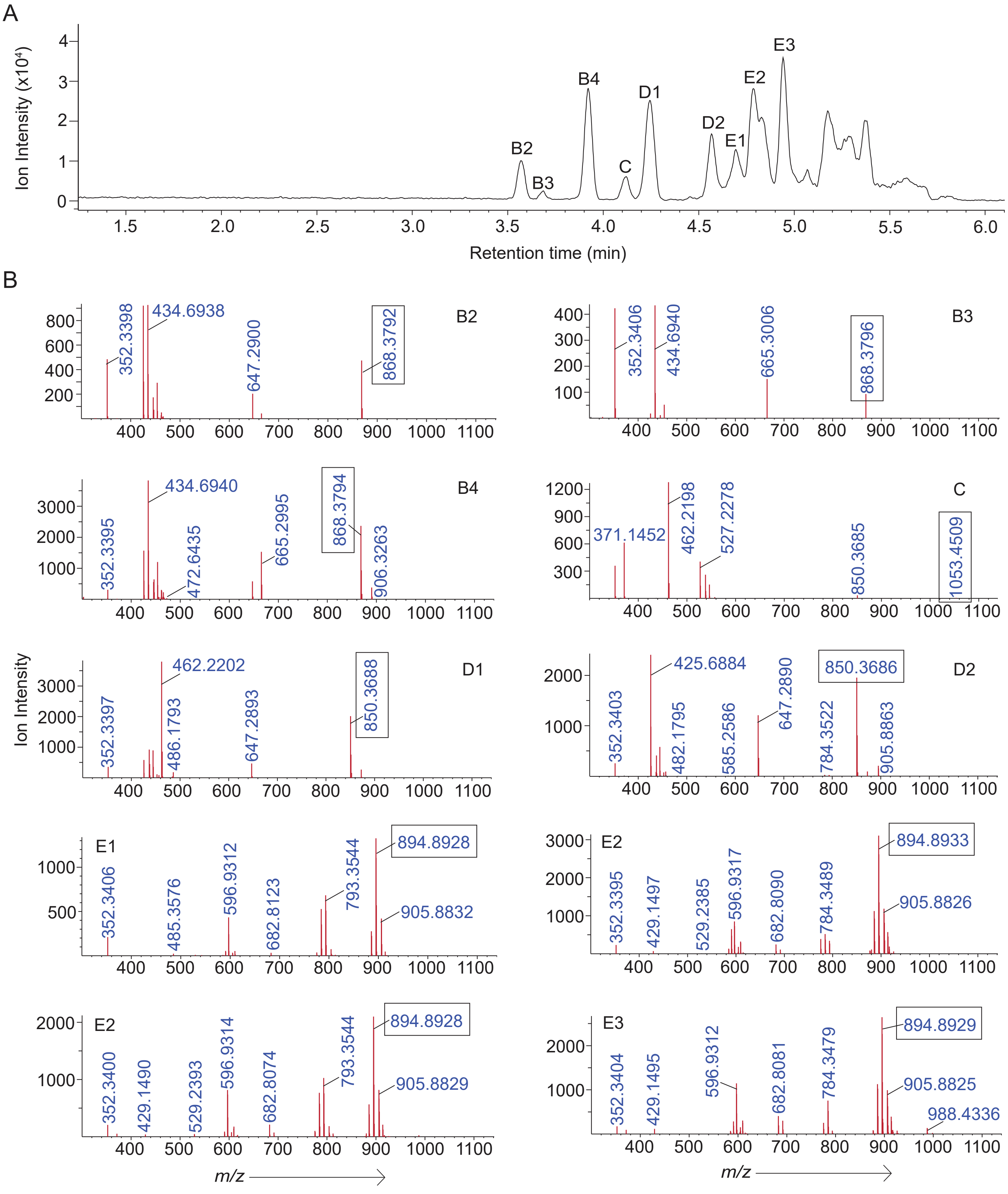


**Fig. S3: LCMS analysis of muropeptides bound to mAb 1A4.** (**A**) LCMS chromatogram showing distinct peaks corresponding to muropeptides captured by 1A4 following incubation with dPG. (**B**) Mass spectra corresponding to each chromatographic peak, confirming the identity of the bound muropeptides. The boxed region highlights the mass spectra peak at the indicated m/z corresponding to the assigned muropeptide. Crosslinked species (structure E), are detected as doubly charged ions (2+), with molecular mass approximated as 2 × m/z.

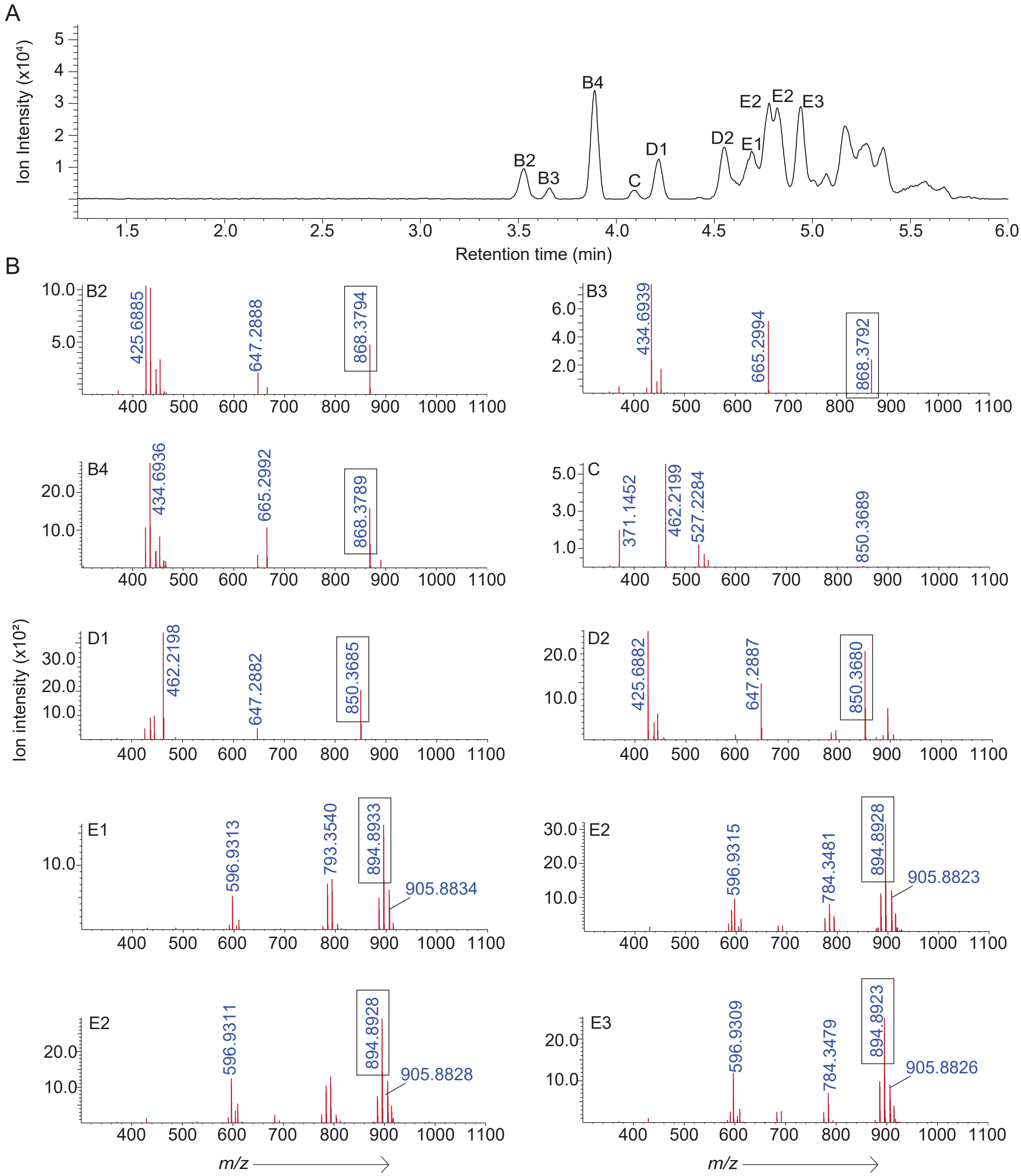


**Fig. S4: LCMS analysis of muropeptides bound to mAb 2H10.** (**A**) LCMS chromatogram showing distinct peaks corresponding to muropeptides captured by 2H10 following incubation with dPG. (**B**) Mass spectra corresponding to each chromatographic peak, confirming the identity of the bound muropeptides. The boxed region highlights the mass spectra peak at the indicated m/z corresponding to the assigned muropeptide. Crosslinked species (structure E), are detected as doubly charged ions (2+), with molecular mass approximated as 2 × m/z.

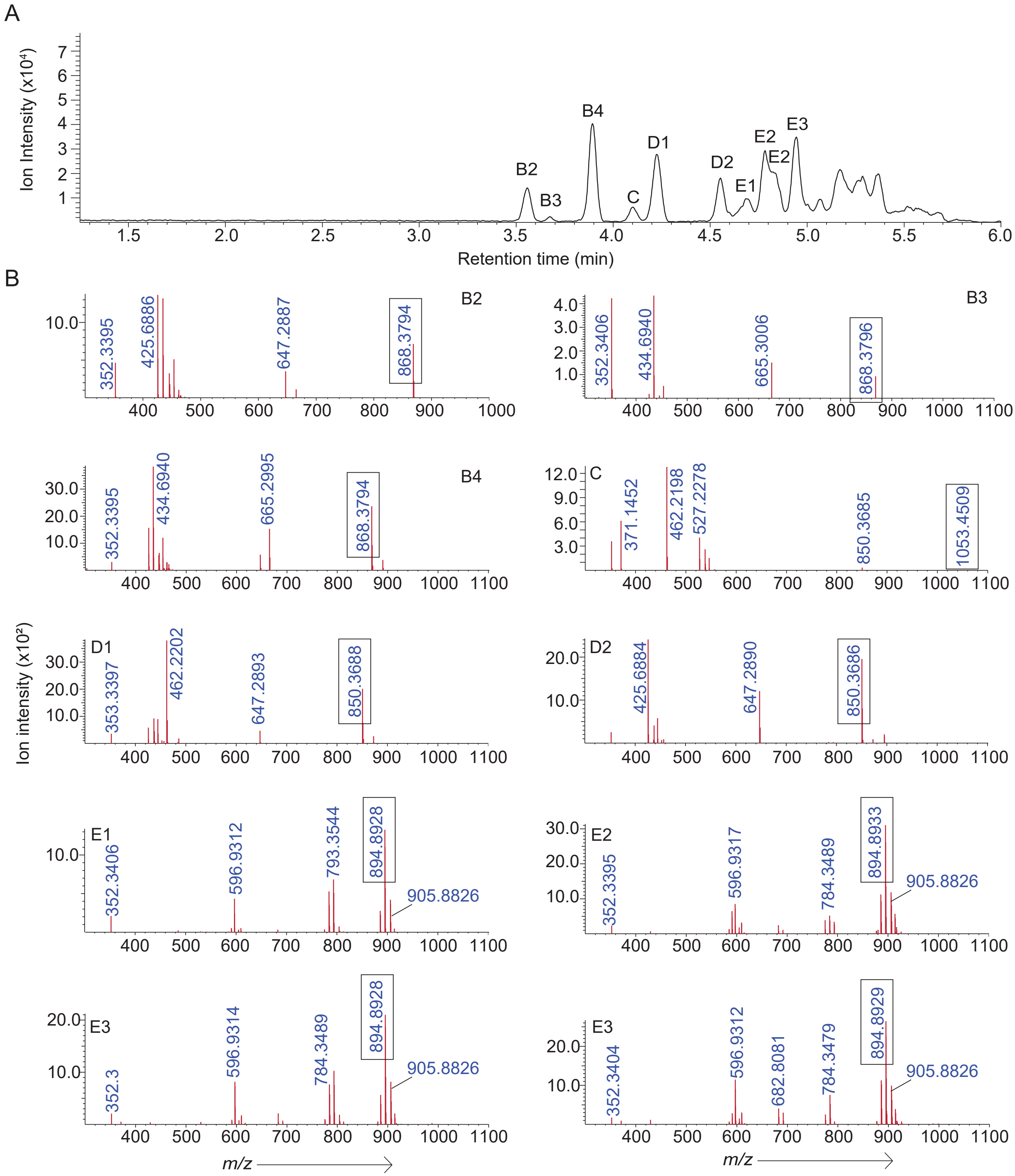


**Fig. S5: LCMS analysis of muropeptides bound to mAb 1B8.** (**A**) LCMS chromatogram showing distinct peaks corresponding to muropeptides captured by 1B8 following incubation with dPG. (**B**) Mass spectra corresponding to each chromatographic peak, confirming the identity of the bound muropeptides. The boxed region highlights the mass spectra peak at the indicated m/z corresponding to the assigned muropeptide. Crosslinked species (structure E), are detected as doubly charged ions (2+), with molecular mass approximated as 2 × m/z.

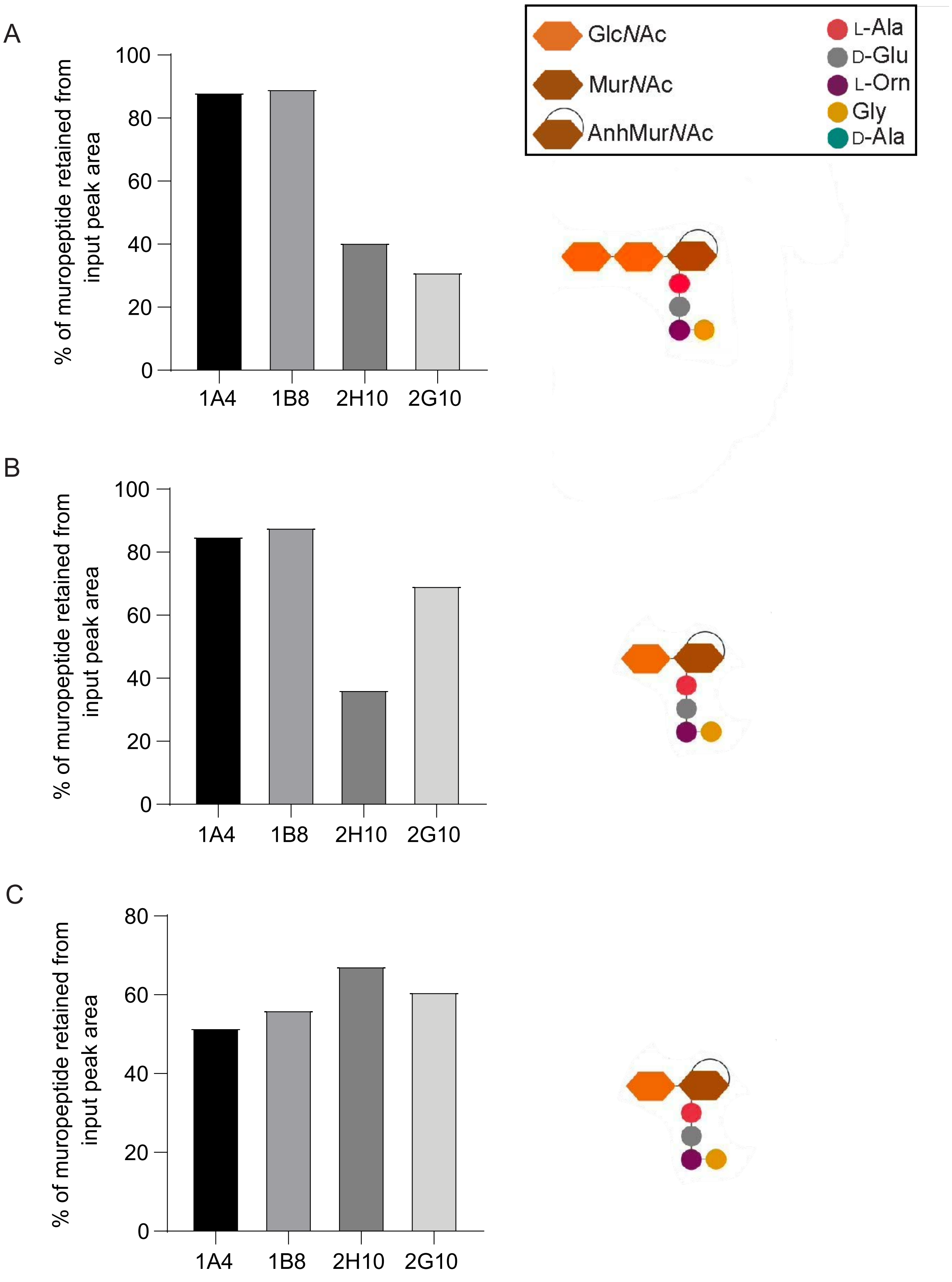


**Fig S6: Muropeptides with a 1,6 anhydromuramic acid group that were identified by LCMS after antibody pull-down.** IP-specificity of each monoclonal for G-G-anhM-AEOG (A) and G-anhM-AEOG (B and C). Results represent the percentage of each muropeptide bound by the four monoclonal antibodies, relative to the input control. Analysis of LCMS data (Figs. S2-S5) identified the structure of each, shown as a cartoon with the corresponding legend shown above (boxed). Note that both 850 species (B and C) are isobaric structures.

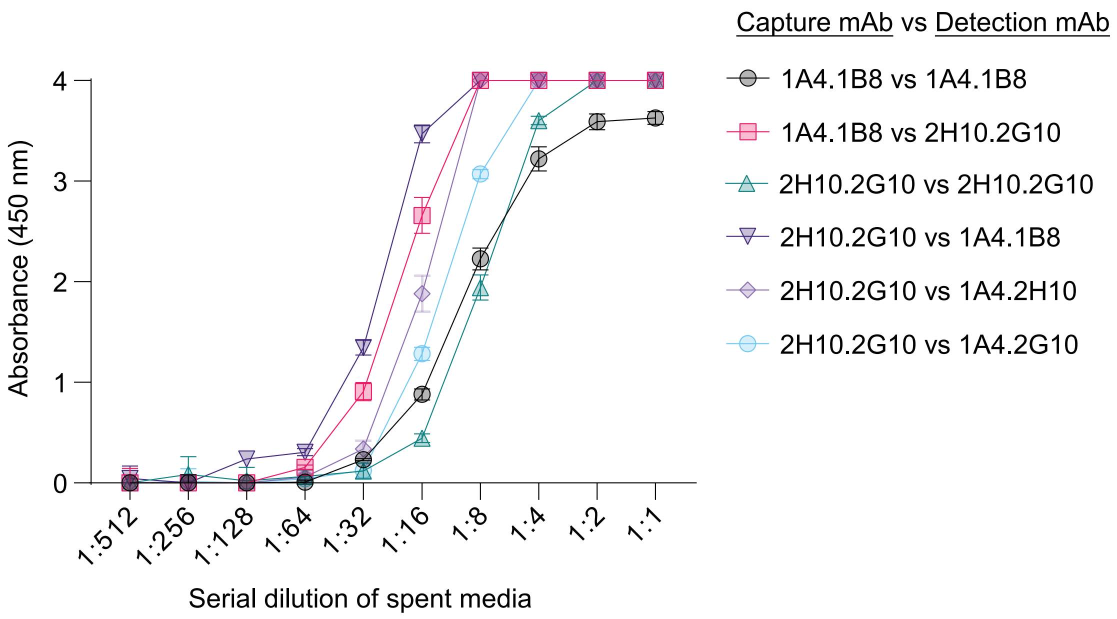


**Fig. S7: Optimization of mAb cocktail to detect released peptidoglycan in *B. burgdorferi* spent culture medium.** Equal amounts of selected monoclonal antibodies were combined and used as the coating antibody. An equivalent mixture of biotinylated monoclonal antibodies served as the detection antibody to identify the combination with the highest affinity for released *B. burgdorferi* PG. Shown are the mean (+/- SD) of triplicate values. Data points with absorbance of 4 were beyond the detection limit of the spectrophotometer.

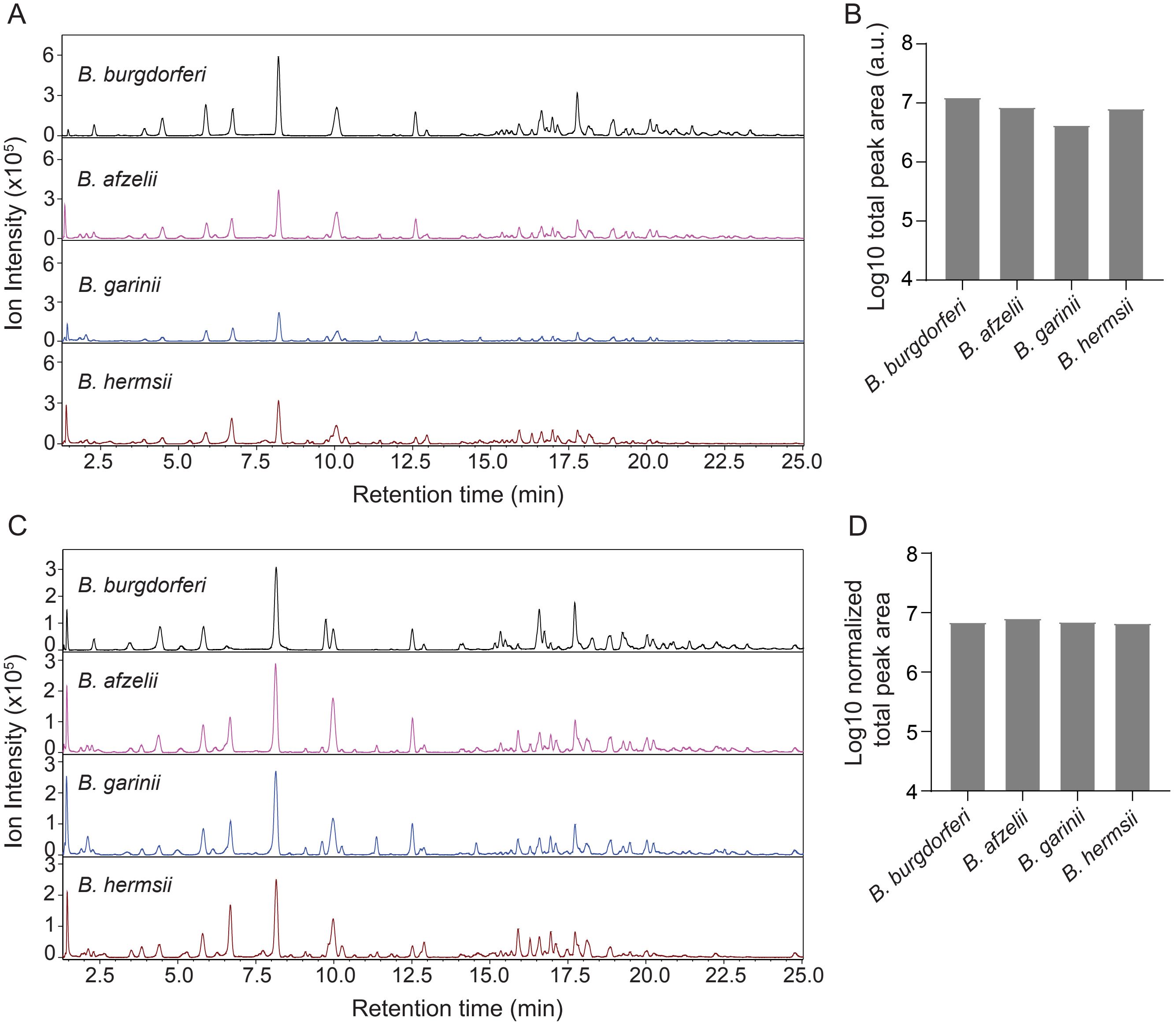


**Fig. S8: LCMS-based normalization of *Borrelia spp*. muropeptides.** PG from multiple *Borrelia spp*. was isolated, enzymatically digested, and analyzed by LCMS to identify muropeptide peaks and quantify total peak area. The summed peak area was used to normalize the amount input PG. After normalization, adjusted amount of PG were re-analyzed by LCMS analysis. **(A)** LCMS chromatograms of PG samples before normalization showing total muropeptide peak signals. **(B)** Quantification of total muropeptide peak area for each sample prior to normalization. **(C)** LCMS chromatograms of PG samples following normalization. **(D)** Quantification of total muropeptide peak area after normalization.

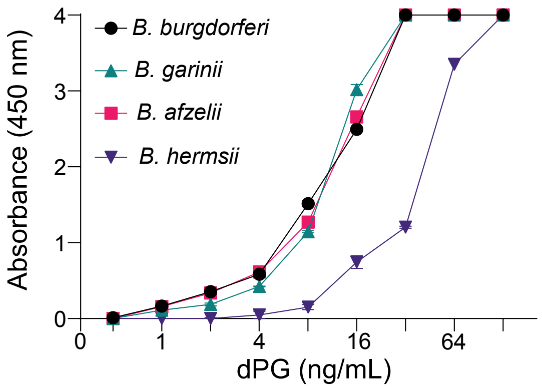


**Fig. S9: Optimized mAb cocktail cross-reacts with digested PG from different Lyme-disease causing *Borrelia spp*.** Purified, dPG attained from different *Borrelia spp*. was titrated in PBS and detected by ELISA. Values indicate the mean (+/- SD) of triplicate samples. Data points with absorbance of 4 were beyond the detection limit of the spectrophotometer.

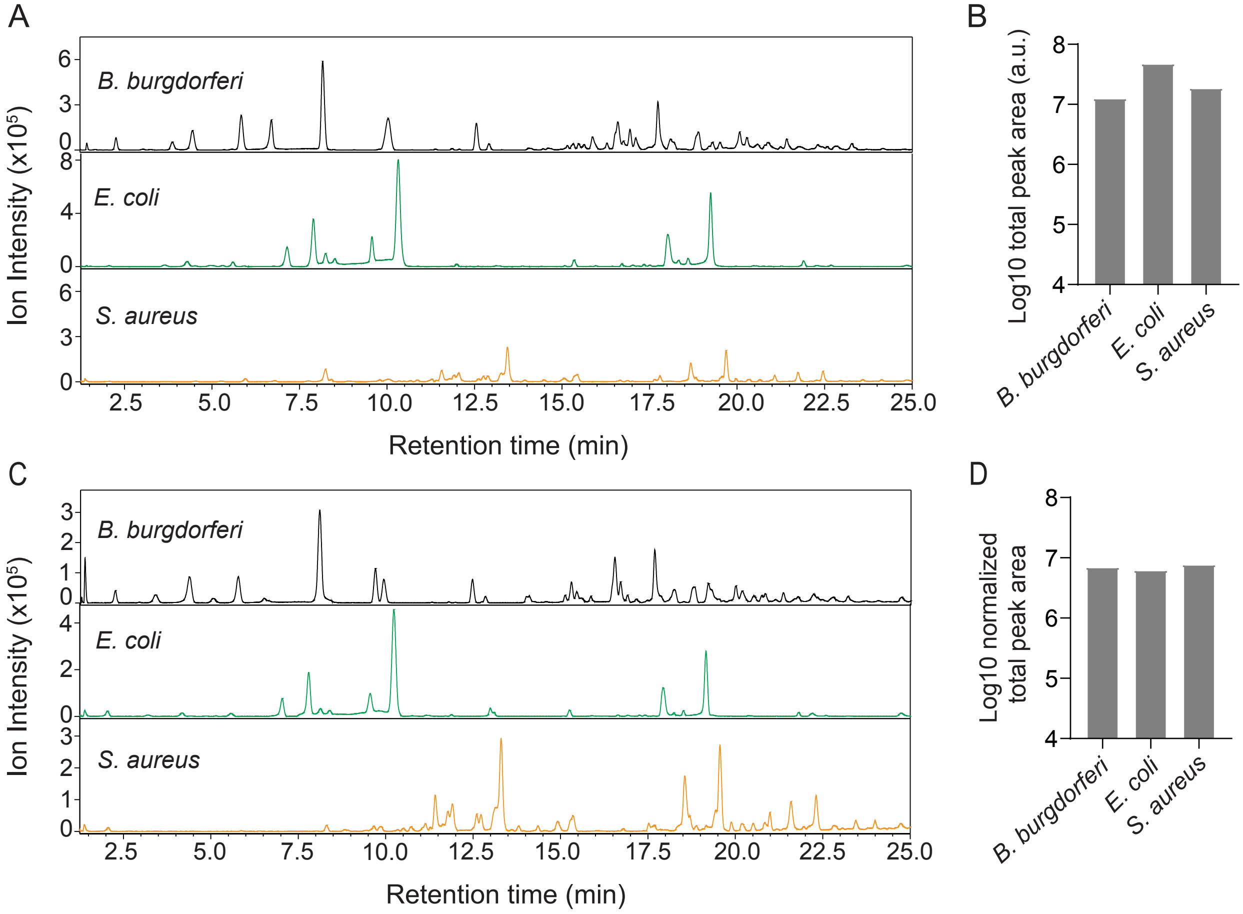


**Fig. S10: LCMS-based normalization of muropeptides across different bacterial *spp.*** PG from *B. burgdorferi, E. coli* and *S. aureus* were isolated, enzymatically digested, and analyzed by LCMS to identify muropeptide peaks and quantify total peak area. The summed peak area was used to normalize the PG input across samples. After normalization, LCMS analysis was repeated. **(A)** LCMS chromatograms of PG samples before normalization showing total muropeptide peak signals. **(B)** Quantification of total muropeptide peak area for each sample prior to normalization. **(C)** LCMS chromatograms of PG samples following normalization. **(D)** Quantification of total muropeptide peak area after normalization.

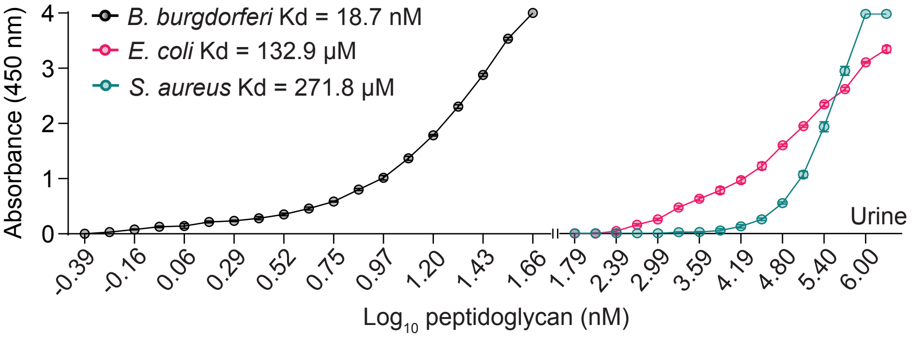


**Fig. S11: Disassociation constant calculations for mAb capture/detection pair**. Sandwich ELISA was used to determine the apparent dissociation constants (Kd) of the mAb cocktail for dPG purified from *B. burgdorferi*, *E. coli*, and *S. aureus* spiked into neat human urine.

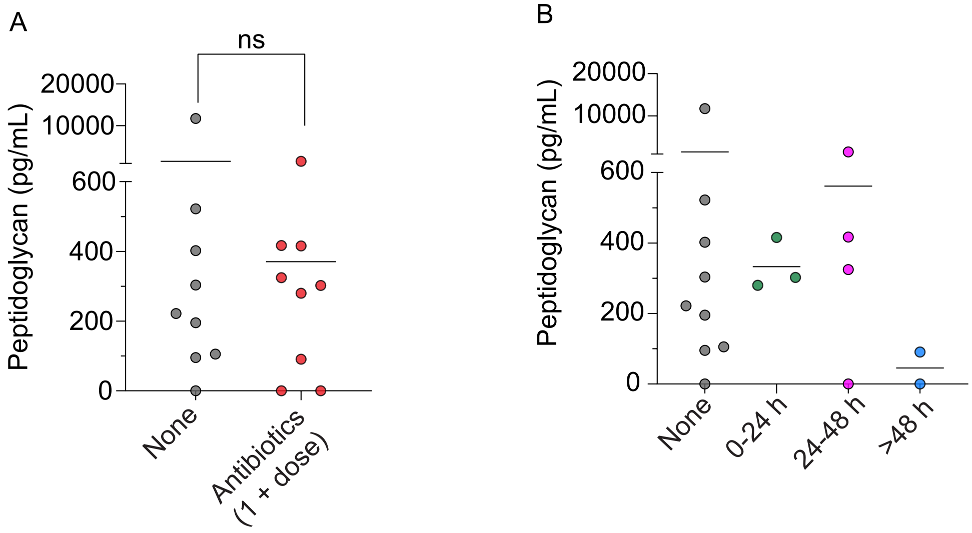


**Fig. S12: Antibiotics treatment status of patients relative to PG levels in urine.** Prior to study enrolment, some patients visited their primary care physician and started doxycycline therapy. **(A)** PG urine levels in patients who received no antibiotics (mean PG 1507 pg/ml) or at least 1 dose (mean PG 370 pg/ml). n.s., not significant by (Mann Whitney U). **(B)** The same data in 'A' approximately straitified based on time/amount of treatment.

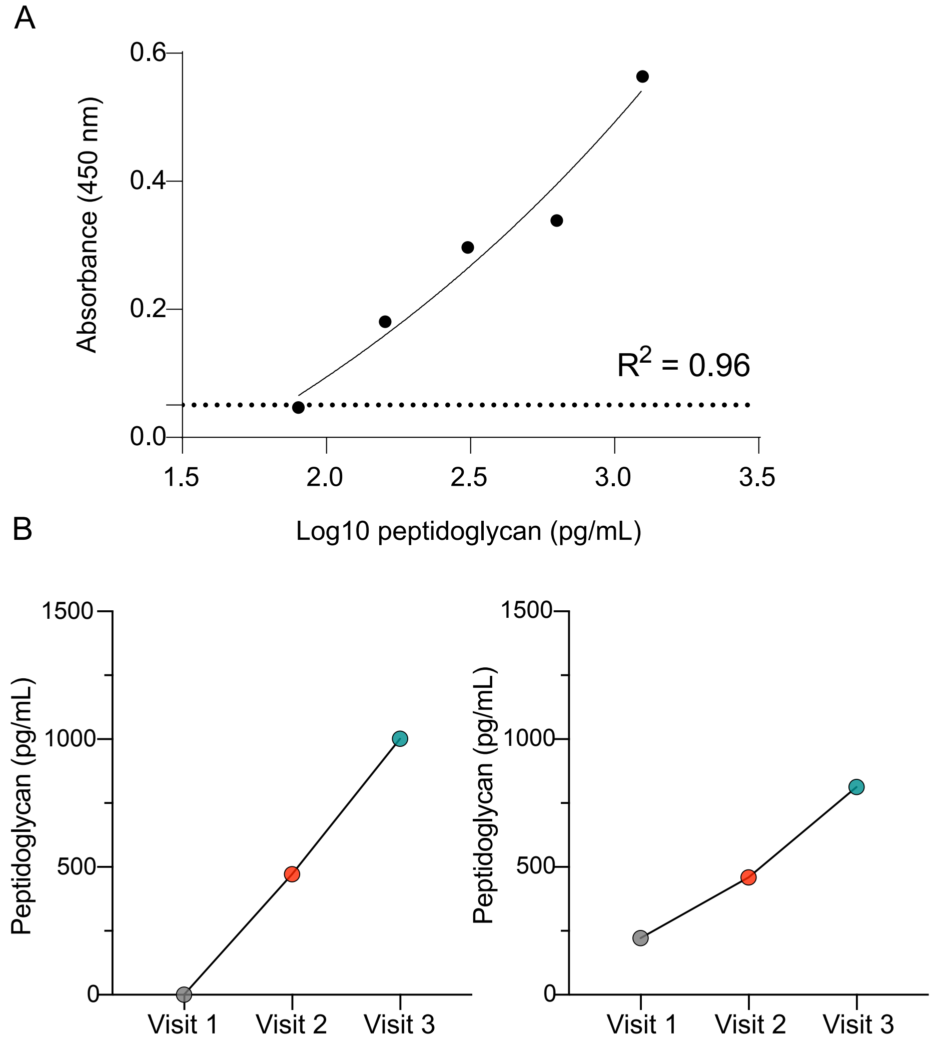


**Fig. S13: Quantifying *B. burgdorferi* peptidoglycan concentration in the urine of human Lyme disease patients.** (**A**) Standard curve generated after background subtraction and used to interpolate PG concentrations in urine samples from human Lyme disease patients. The horizontal black line indicates the limit of detection, defined as 3× the standard deviation of the blank. Samples with absorbance values at or below this threshold (≤ LOD) were considered undetectable for PG. (**B**) Longitudinal measurements of PG levels in patient samples by sandwich ELISA at baseline (visit 1), 30 days (visit 2), and 90 days (visit 3) from the two individuals (left and right) who contained detectable amounts after treatment.
